# Reconstructing the force of infection and immune fraction of the population via a single snapshot survey: a case study of COVID-19 in Japan

**DOI:** 10.1101/2025.04.08.25325449

**Authors:** Yuta Okada, Hiroshi Nishiura

**Author notes:** **Correspondence to**: H. Nishiura, Kyoto University School of Public Health, Yoshida-Konoe, Sakyo, Kyoto 606-8601, Japan.

## Abstract

While the global health burden of COVID-19 continues, multifaceted epidemiological surveillance is required to monitor the epidemic’s dynamics and its population-wide risk. We proposed a simple framework using a population-wide snapshot questionnaire survey to estimate the incidence and protective effect of immunity by natural infection or vaccination against the SARS-CoV-2 JN.1 variant.

Our results revealed that in Japan in February 2024, the personal risk of infection was substantially higher in younger adults and risk was heterogenous across prefectures. Diabetes mellitus (relative risk 1.8; 95% credible interval [CrI] 1.1, 2.9), neoplastic disorders (5.2; 95% CrI 3.1, 8.6), immunological suppression (2.6; 95% CrI 1.3, 4.6), respiratory diseases (2.2; 95% CrI 1.46, 3.3), and cardiovascular disease (2.3; 95% CrI 1.3, 3.9) were risk factors for infection.

The highest peak protection after infection was after exposure to pre-XBB.1.5 Omicron variants (52.0%; 95% CrI 33.2, 68.7), whereas the XBB.1.5 monovalent vaccine provided the highest protection (45.1%; 95% CrI 37.8, 52.7) among three vaccine types. Notably, the peak protection of the bivalent Wuhan + Omicron BA.1/5 vaccine was substantially lower than other vaccines (28.7; 95% CrI 17.3, 40.6). By statistical matching the respondent cohort to the 2020 population census, we revealed that the national COVID-19 incidence rate in February 2024 by age group was highest (4.73%; 95% CrI 4.17, 5.38) and lowest (1.19%; 95% CrI 0.94, 1.47) among those aged 20–29 years and 60–69 years, respectively. The force of infection was high and more heterogeneous in younger groups, whereas younger populations were more concentrated than older populations in low-protection regions.

Our framework revealed biological and epidemiological insights into protection and risk of infection from past immunizing events and personal attributes during the JN.1-dominant period. Moreover, we proposed a framework for the rapid evaluation of epidemiological dynamics whose application is not limited to COVID-19.

**Author Summary:** The questionnaire survey is a powerful tool for generating population-wide data in fields including public health, but its application in tracking the immunological characteristics and incidence rate of COVID-19 is limited. We proposed a new framework to leverage the advantages of this tool by combining an Internet-based questionnaire survey with statistical models. Two findings emerged from our study, which focused on COVID-19 infection in February 2024, when the JN.1 variants of severe acute respiratory syndrome coronavirus 2 were dominant in Japan. First, the overall reduction in the relative risk of infection from vaccinations or natural infection was at most roughly 60% and had substantial time decay. Second, compared with older age groups, a higher incidence rate and lower protection from acquired immunity were observed in young age groups, with substantial heterogeneity within each age-stratified population. This study revealed important immune profiles against the emerging JN.1 variant and identified an effective framework that can be added to the array of epidemiological programs that complement routine public health surveillance.

## 1. INTRODUCTION

The coronavirus disease 2019 (COVID-19) pandemic has caused extensive mortality since its emergence in 2019 in Wuhan, China [1–3]. Although COVID-19 continues to have a substantial impact on health [4–9], epidemiological surveillance efforts have been globally downgraded to the extent that tracking the absolute number of infections and transmission dynamics at the national and regional levels has become impossible in most countries. Furthermore, at more than 4 years since the emergence of the virus, conducting studies on the effectiveness of vaccine- or infection-induced immunity has become increasingly complex given the growing complexity of the individual-level history and antigenic details of vaccination and infection and the very small size of the population naïve to both infection and vaccination. In addition, adherence by individuals to precautionary measures has varied considerably over time given society’s increasingly diverse views toward COVID-19. Because of the increased number of infections and the accelerated evolution of the virus, the rapid turnover of SARS-CoV-2 variants has also thwarted conventional vaccine studies that quantify protection level by measuring humoral immune response. Given that the results of such studies reach the public domain in a few months, a new SARS-CoV-2 variant with a substantial transmission advantage or immune evasion capability can begin causing another large epidemic. To facilitate the use of population immune profiles as part of real-time epidemiological risk assessment, approaches that expand the coverage and timeliness of epidemiological surveillance are urgently needed.

Efforts to enhance the coverage and timeliness of epidemiological surveillance beyond routine public health monitoring have been made before the emergence of COVID-19. During the COVID-19 pandemic, most countries ran universal or sentinel surveillance to monitor the virus, whereas various complementary approaches such as wastewater surveillance, digital syndromic surveillance, and participatory surveillance, including population-wide questionnaire surveys, increasingly proved useful during the pandemic [10–19]. These approaches provide tools to capture the epidemiological trend of the epidemic. However, to capture the entire scale of an epidemic, participatory or questionnaire-based survey can also be practical options. Cross-sectionally, a participatory active survey involving biological samples has been proven valuable [19], although financial and labor costs may limit its deployment. Additionally, although pure questionnaire surveys cannot collect biological evidence, they can 1) play an important role in quantifying transmission dynamics, 2) be implemented at a low cost, and 3) be conducted rapidly. Published studies have deployed questionnaire surveys together with epidemiological indicators such as the case-fatality ratio to estimate the population-wide incidence of influenza in the United States [18,20]. These studies were based on telephone surveys; Internet-based surveys can further enhance the effectiveness of questionnaire surveys in the context of epidemic risk assessment.

Studies in the earlier COVID-19 pandemic period that estimated the immunity effects of infection or vaccination after adjustment for individual factors, including health, biological, and social status, have been extremely successful [21,22]. However, because the variability of personal precautionary behaviors is currently high, regularly updating knowledge on the effect of factors that modulate the susceptibility to infection is desirable. Furthermore, providing national or sub-population-level “susceptibility mapping” by attributes such as age can be invaluable for guiding public health decisions that optimize non-pharmaceutical interventions or vaccination strategies for epidemic control [23]. Published studies have provided useful frameworks for susceptibility mapping; however, these frameworks do not apply to the current global situation in which population-level data on case counts and vaccination records are no longer available [24,25].

In Japan, the universal COVID-19 case count was officially stopped in May 2023, and national vaccination records (e.g., as a registration system) are also currently unavailable. Some projects are underway to recover, at least partially, case counts before May 2023 that are equivalent to universal case counts, and a government-led project is underway to conduct seroepidemiological surveys among voluntary participants and collect residual serum samples from healthcare facilities and blood donors [10,26,27]. However, although these efforts are very useful for addressing the current lack of comprehensive epidemiological data, the coverage of these projects is insufficient for rapidly elucidating population-level incidence and immunity. Moreover, at the time of writing, vaccine effectiveness in Japan against infection or severe disease from COVID-19 had only been evaluated up to the bivalent mRNA vaccine and for COVID-19 variants that preceded the JN.1 variants [28–33]. Thus, identifying a framework that enables timely updates of personal-level immunity against the latest emerging SARS-CoV-2 variants is another important goal. In this context, we propose a web-based questionnaire approach to estimate not only personal-level immunity and risk-modifying attributes, including health conditions, but also the population-level incidence of COVID-19 infection and immune protection.

Motivated by the issues described above, we proposed a practical framework that uses an Internet-based social survey to estimate the COVID-19 incidence rate and immune protection against the virus from past infection or vaccination. Using survey respondents’ personal background information, we scaled up the survey results to reconstruct the population-wide incidence and immune profiles. This process was achievable only via a single snapshot survey.

## 2. RESULTS

### 2.1 Force of infection and relative risk by personal attribute

Table 1 summarizes the posterior estimates of *β*_*k*_. The intercept *β*_0_ represents the baseline national force of infection (FoI) among those aged 20–29 years without any additional effects related to personal attributes. An inverse relationship between the relative risk of infection and age was observed. Among health condition factors, substantially high relative risks of infection were observed in diabetes mellitus (1.8; 95% CrI 1.1, 2.9), neoplastic disorders (5.2; 95% CrI 3.1, 8.6), immunological suppression (2.6; 95% CrI 1.3, 4.6), respiratory disease (2.2; 95% CrI 1.4, 3.3), and cardiovascular disease (2.3; 95% CrI 1.3, 3.9). A similar trend was suggested in liver disease. Obesity did not substantially modulate the relative risk of infection (0.7; 95% CrI 0.3, 1.3). Additionally, smoking (1.3; 95% CrI 1.0, 1.8) and alcohol drinking (0.9; 95% CrI 0.6, 1.3) did not significantly alter the risk of infection. A previous history of infection (7.7; 95% CrI 5.6, 10.7) was strongly associated with the risk of infection. Although not shown in Table 1, prefectural analysis revealed a mildly higher risk in specific prefectures, including Ishikawa, Osaka, and Kagoshima (S1 Fig).

**Table 1.** Estimates of relative risk of infection by personal attribute.

| Variable | Estimated values (95% CrI) |
| --- | --- |
| Intercept | 0.017 (0.011, 0.025) |
| age group 30-39 (years) | 1.040 (0.766, 1.447) |
| age group 40-49 (years) | 0.768 (0.528, 1.092) |
| age group 50-59 (years) | 0.556 (0.352, 0.867) |
| age group 60-69 (years) | 0.372 (0.214, 0.609) |
| age group 70 years and older | 0.367 (0.204, 0.608) |
| Female Sex (dichotomous) | 0.851 (0.657, 1.084) |
| Diabetes Mellitus | 1.832 (1.107, 2.949) |
| Neoplastic Disorders | 5.249 (3.130, 8.563) |
| Immunologically Suppressed | 2.599 (1.337, 4.558) |
| Respiratory Diseases | 2.174 (1.361, 3.342) |
| Cardiovascular Diseases | 2.279 (1.262, 3.910) |
| Cerebrovascular Diseases | 1.503 (0.756, 3.305) |
| Liver Disorders | 1.911 (0.957, 3.717) |
| Obesity (BMI >30kg/m<sup>2</sup>) | 0.732 (0.339, 1.273) |
| Smoking habit | 1.306 (0.950, 1.797) |
| Alcohol drinking habit | 0.900 (0.589, 1.276) |
| Household size $\geq 2$ | 0.847 (0.631, 1.127) |
| History of Previous Infection | 7.687 (5.608, 10.70) |

### 2.2. Protection against COVID-19 infection from infection or vaccination in February 2024

The protection or relative risk reduction against COVID-19 from natural infection or vaccination is summarized in Fig 1. At the peak of protective immunity (i.e., at 30 days after the last immunizing event), protection levels provided by infections were higher overall than those by vaccines. At 30 days post-infection, the highest protection level (52.0% risk reduction; 95% CrI 33.2, 68.7) was against pre-XBB Omicron variants, followed by that against XBB.1.5 lineages (47.2%; 95% CrI 32.3, 60.7) and pre-Omicron variants (42.4%; 95% CrI 24.2, 50.5). At 30 days post-vaccination, the protection offered by the XBB.1.5 monovalent vaccine (45.1%; 95% CrI 37.8, 52.7) was the highest, followed by that of the Wuhan monovalent vaccine (36.3%; 95% CrI 19.8, 54.1) and the Wuhan-Omicron BA.1/5 bivalent vaccine (28.7%; 95% CrI 17.3, 40.6). The parameters underlying these estimates are provided in Table 2.

**Table 2.** Estimated Peak protection (expressed as relative risk reduction in percentage) at one month after the last immunizing event by type of vaccine or natural infection.

| Last Immunizing Event | Estimated values (95% CrI) |
| --- | --- |
| Infection |  |
| XBB sub lineages | 47.2 (32.3, 60.7) |
| pre-XBB Omicron | 52.0 (33.2, 68.7) |
| pre-Omicron | 42.4 (24.2, 60.5) |
| Vaccination |  |
| XBB.1.5 | 45.1 (37.8, 52.7) |
| Wuhan + Omicron BA.1/5 | 28.7 (17.3, 40.6) |
| Wuhan | 36.3 (19.8, 54.1) |

**Fig 1.**
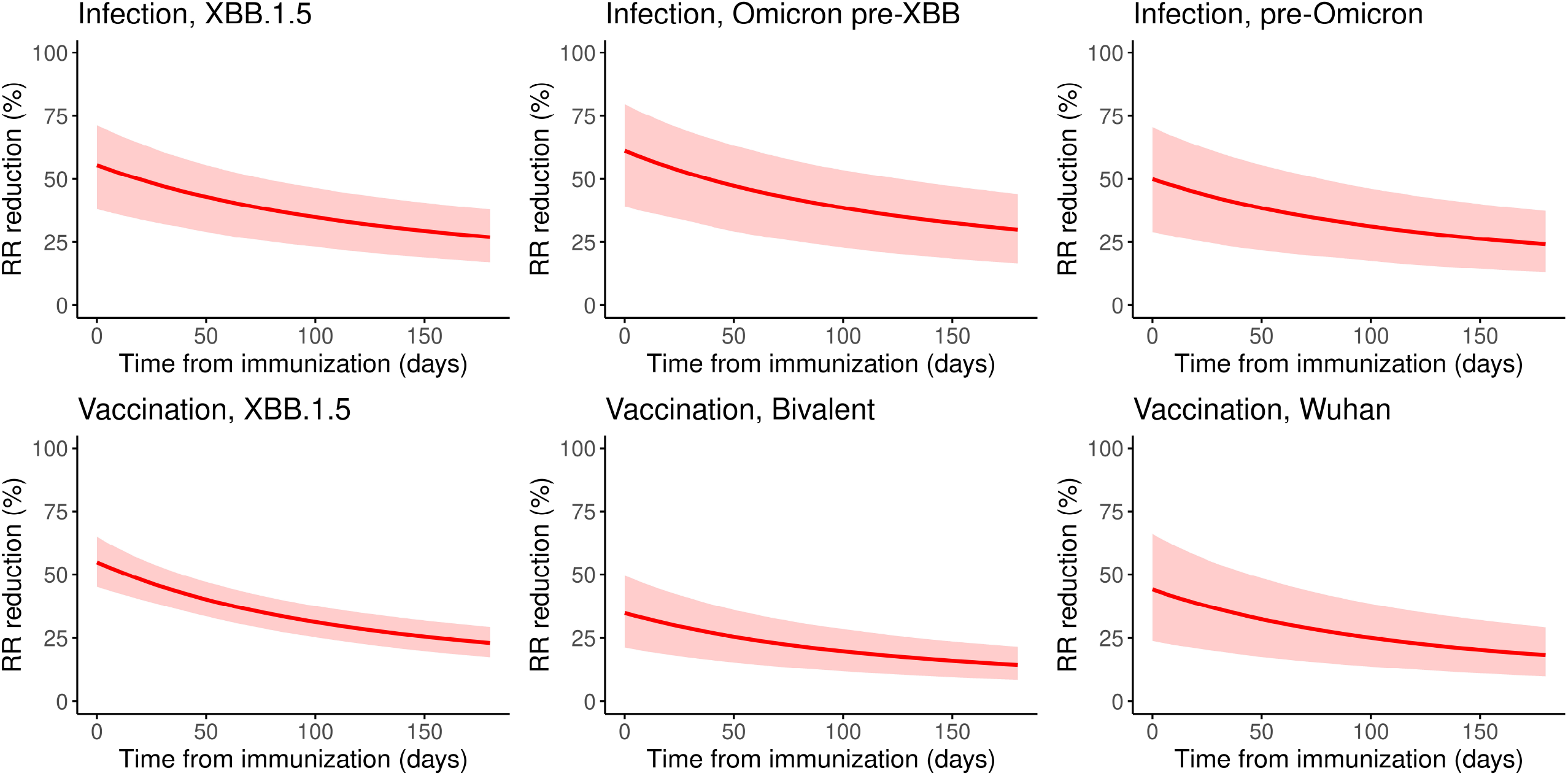
Model estimates of protection levels and their decay over time by type of last immunizing event. Each panel represents the decay of protective immunity (expressed as relative risk reduction) by type of last immunizing event. In each panel, the horizontal axis shows the time since the last immunizing event.

### 2.3. Statistical fitting of the respondent population to the census data

As described in the Materials and Methods section, we estimated the statistical weights for respondents that are needed to convert the result from the respondent group to that equivalent to the population in the 2020 census. Table 3 summarizes the following statistics, which were estimated as the weighted sum of answers from respondents: 1) proportions of men and women in the national adult population, 2) proportions by age group in the national male population, and 3) proportions by age group in the national female population. The proportion of men was estimated at 49.37% (95% CrI 49.36, 49.38), compared with 49.41% based on the 2020 census. Among the male population, the estimated proportions of the population for the 20–29, 30– 39, 40–49, 50–59, 60–69, and 70-year-and-older age groups were 13.43% (95% CrI 13.42, 13.44), 15.52% (95% CrI 15.51, 15.53), 20.09% (95% CrI 20.07, 20.10), 18.04% (95% CrI 18.03, 18.05), 16.61% (95% CrI 16.60, 16.62), and 16.31% (95% CrI 16.30, 16.32), respectively. Except for the slight discrepancies in the 20–29-year and 70-year-and-older age groups, these results mostly aligned with the 2020 census. The estimated age-specific proportions of the female population for the same age groups were 12.72% (95% CrI 12.71, 12.73), 14.73% (95% CrI 14.72, 14.74), 19.24% (95% CrI 19.23, 19.25), 17.67% (95% CrI 17.66, 17.68), 17.03% (95% CrI 17.02, 17.04), and 18.62% (95% CrI 18.60, 18.63), respectively; these results were in line with the 2020 census. The estimated population distribution by prefecture and job type distribution by age group are in S1-S4 Data. Despite slight discrepancies between the 95% CrI and the census data (especially in data categories with sparse absolute counts in the respondent group), the weighted demographics closely followed the pattern in the census data.

**Table 3.**
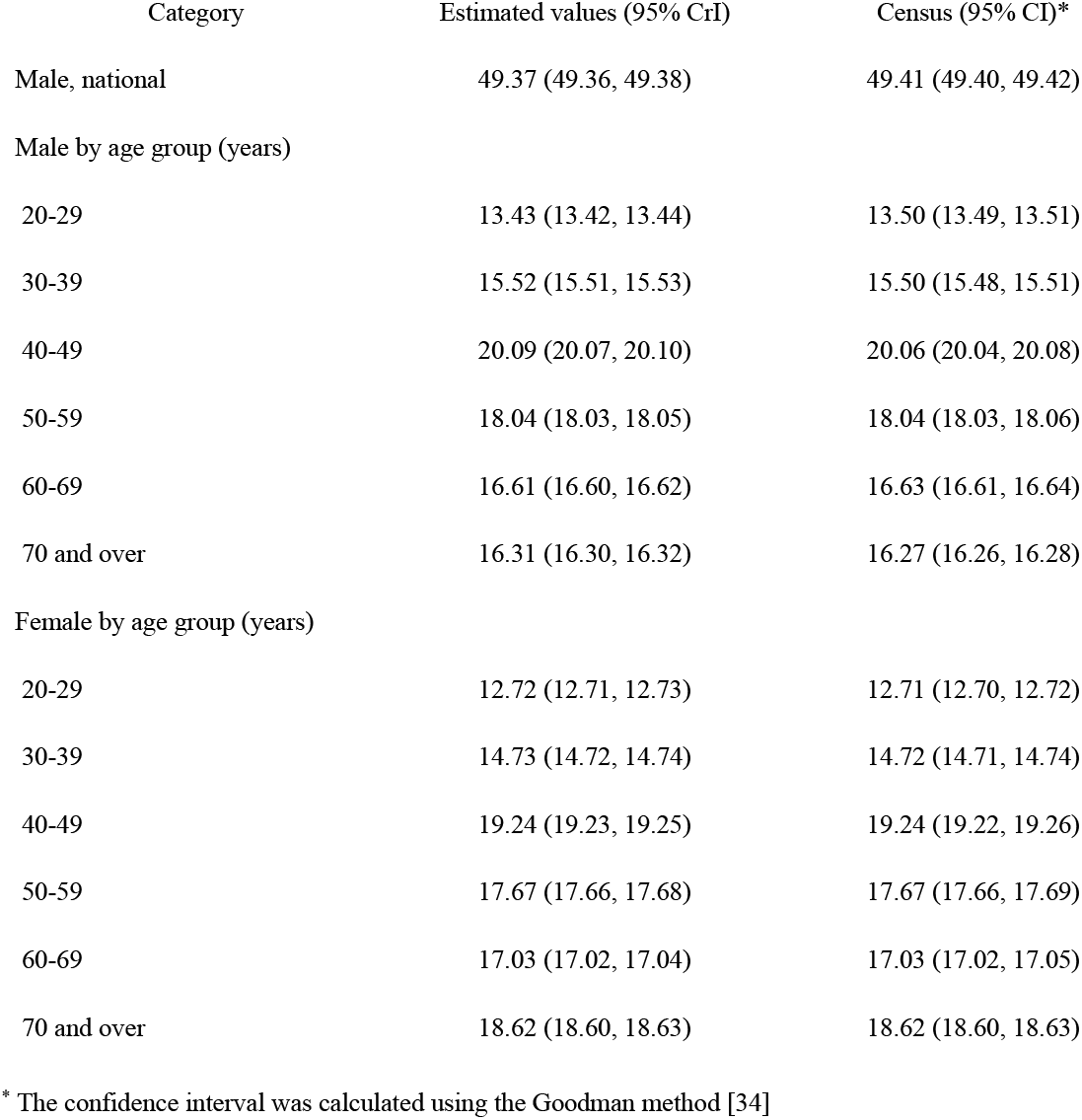
Estimated sex ratio and proportion of population by age group.

### 2.3. COVID-19 incidence by age group in February 2024

Fig 2 shows the estimated COVID-19 incidence rates in the 20–29, 30–39, 40–49, 50–59, 60–69, and 70-year-and-older age groups. The highest estimated incidence was 4.75% (95% CrI; 4.18, 5.40) in those aged 20–29 years, whereas the lowest estimated incidence was 1.18% (95% CrI; 0.94, 1.48) in the 60–69-year age group. Overall, the incidence rates were high in the younger population and low in the older population, with the exception of a higher incidence in the 70-year-and-older age group compared with the 60–69-year age group.

**Fig 2.**
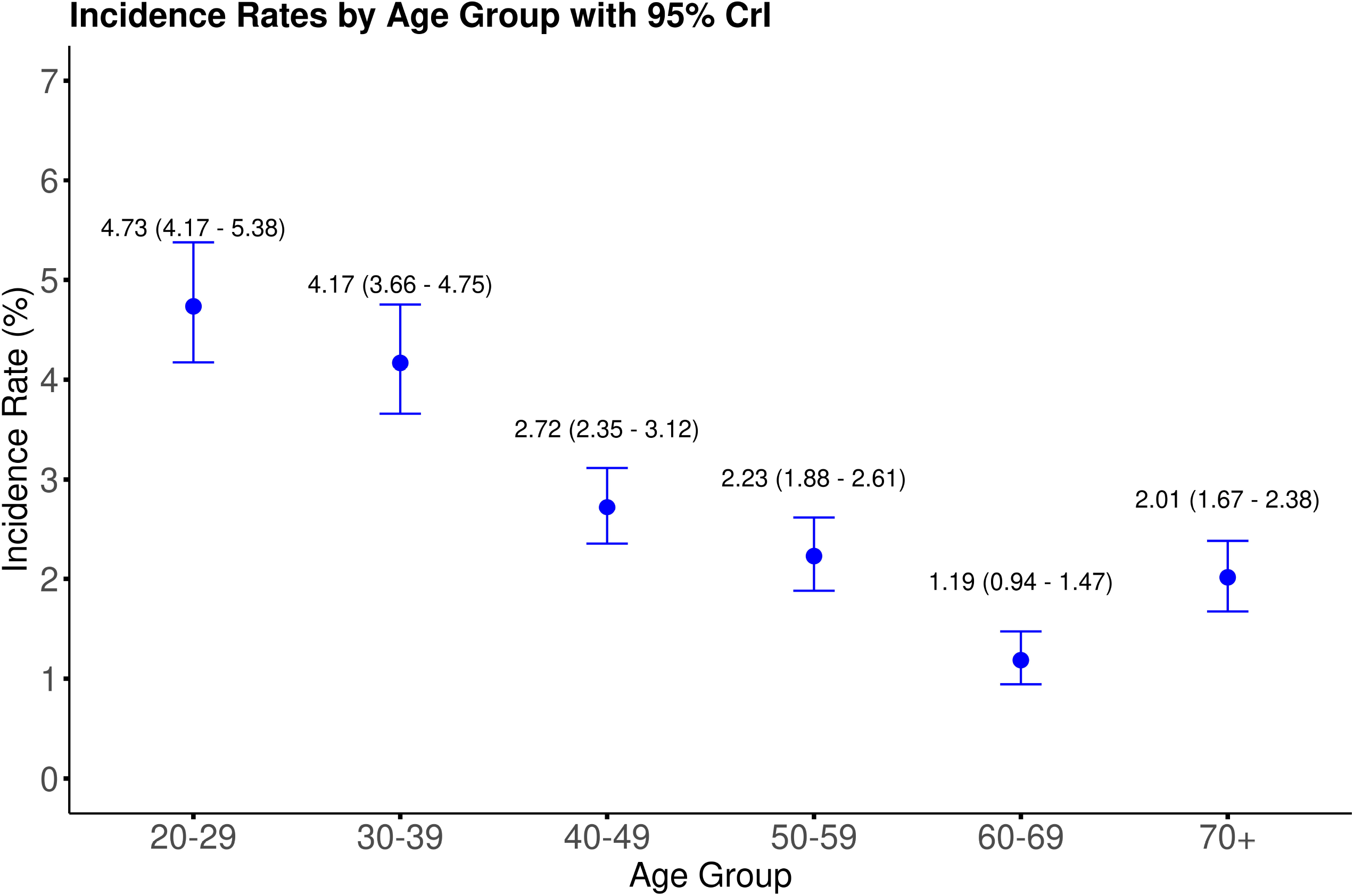
Estimated national incidence rates of COVID-19 by age group in February 2024. For each age group, the blue point represents the median posterior incidence rate estimate, and the error bar represents the 95% credible interval.

### 2.4. Mapping the force of infection and immune protection by age group

Fig 3 illustrates the FoI distribution by each age group. The distributions were created by adapting the statistical weights of respondents by adjusting the questionnaire-based estimates to those equivalent to the national population (See Materials and Methods and S2 Text for further details on the statistical weights). Among those aged 20–29 years, a mild bimodal pattern in the FoI distribution was observed, with over 50% of the population concentrated in the low FoI range (< 0.02) and a substantial proportion also observed in the high FoI range (approximately 0.06–0.14). A very small fraction with FoI > 0.2 was also observed as a slight upward tick in the right tail of the distribution. In older age groups, the distribution was more heavily concentrated in the low FoI range. The bimodal pattern weakened in older ages and was visually unidentifiable among those aged 70 years and older.

**Fig 3.**
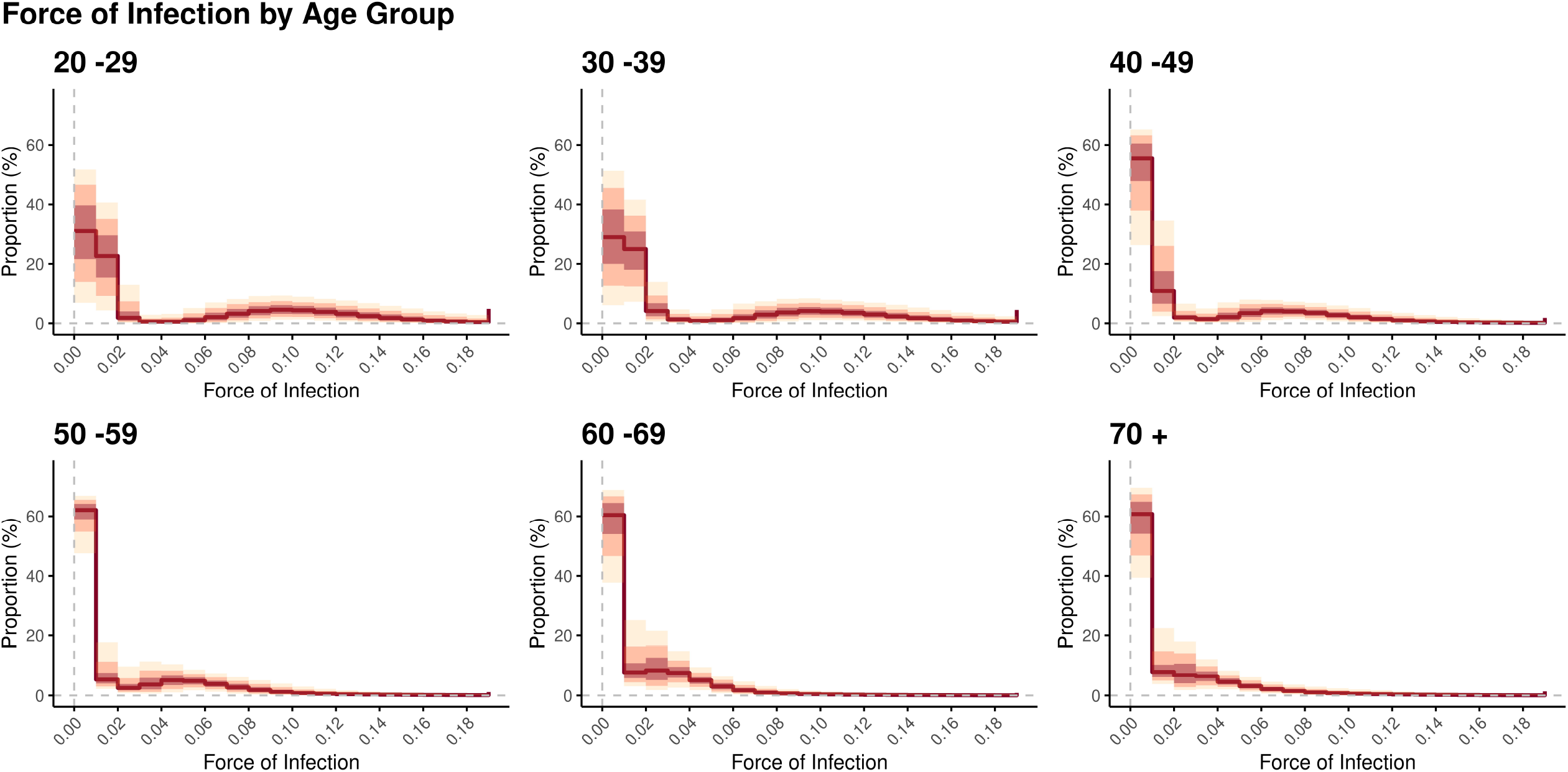
Estimated distribution of the national force of infection by age group in February 2024. Each panel shows the fraction of the age-stratified population that lies in discretized intervals of the force of infection (expressed with the unit of per month). In each panel, the red solid line represents the median in each interval, and the red, pink, and yellow bands represent the 50%, 80%, and 95% credible intervals, respectively.

Fig 4 illustrates the distribution of individual protection as relative risk reduction in each age group. The statistical weights of respondents were adapted in the same manner as in Fig 3. Most of the individuals aged 20–29 years were concentrated in the low protection zone (i.e., below 30%), and a monotonic decline toward the higher protection levels was observed. This pattern weakened in older age groups in which increasingly bimodal distributions of protection levels were observed.

**Fig 4.**
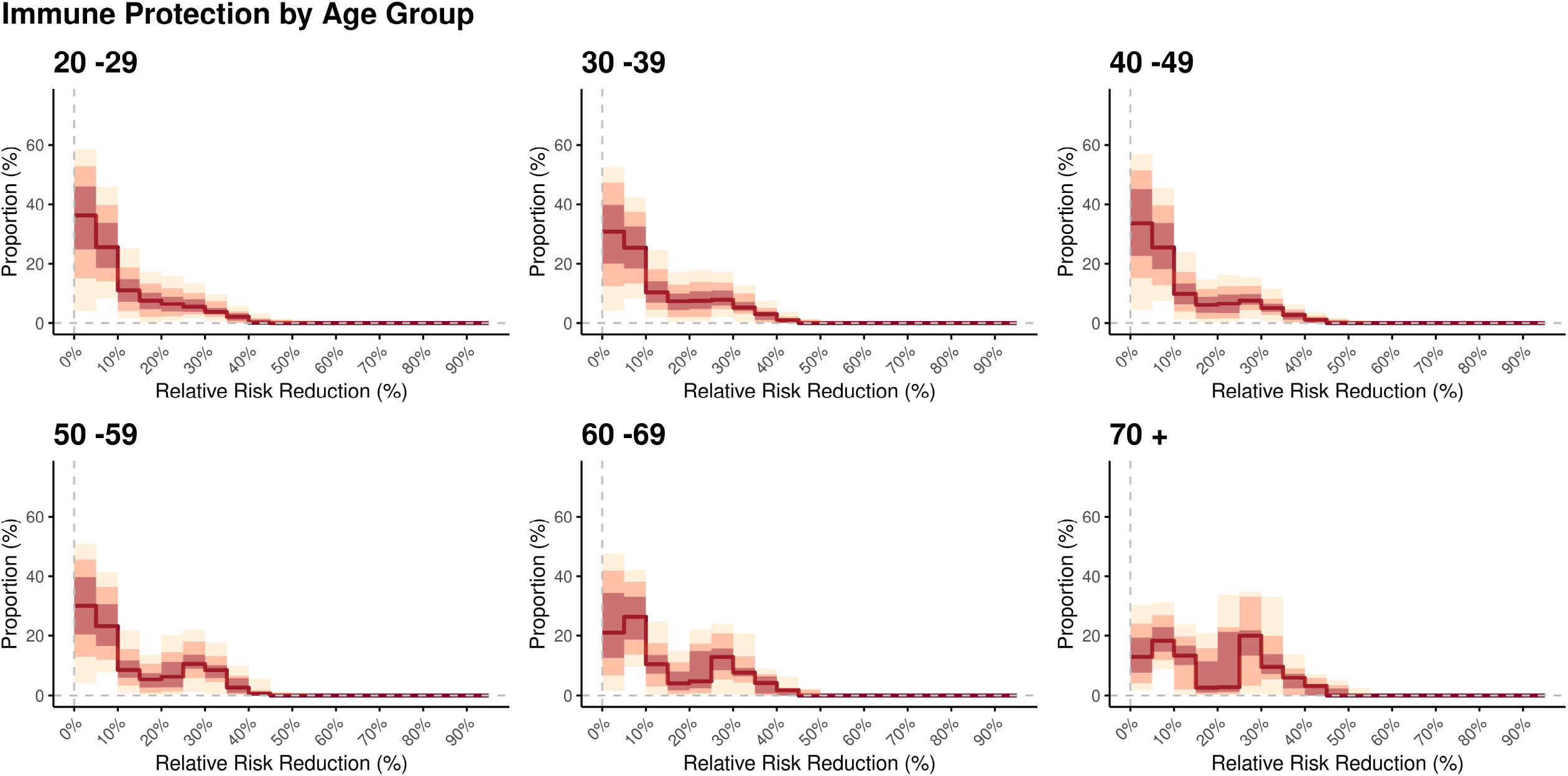
Estimated distribution of national immune protection (expressed as relative risk reduction) by age group in February 2024. Each panel shows the fraction of the age-stratified population that lies in discretized intervals of the protection level. In each panel, the red solid line represents the median in each interval, and the red, pink, and yellow bands represent the 50%, 80%, and 95% credible intervals, respectively.

The findings in Figs 3 and 4 are illustrated as two-dimensional heatmaps in Fig 5, in which the colors in each cell represent the value of the posterior average. In addition to the concentration in the low protection–low FoI region, a substantial fraction in the relatively high FoI range that was centered at roughly 0.10 was observed in younger age groups. This pattern was less evident in the groups aged 60–69 years and 70 years and older. The fraction of those with FoI > 0.2 also tended to decrease in older age groups.

**Fig 5.**
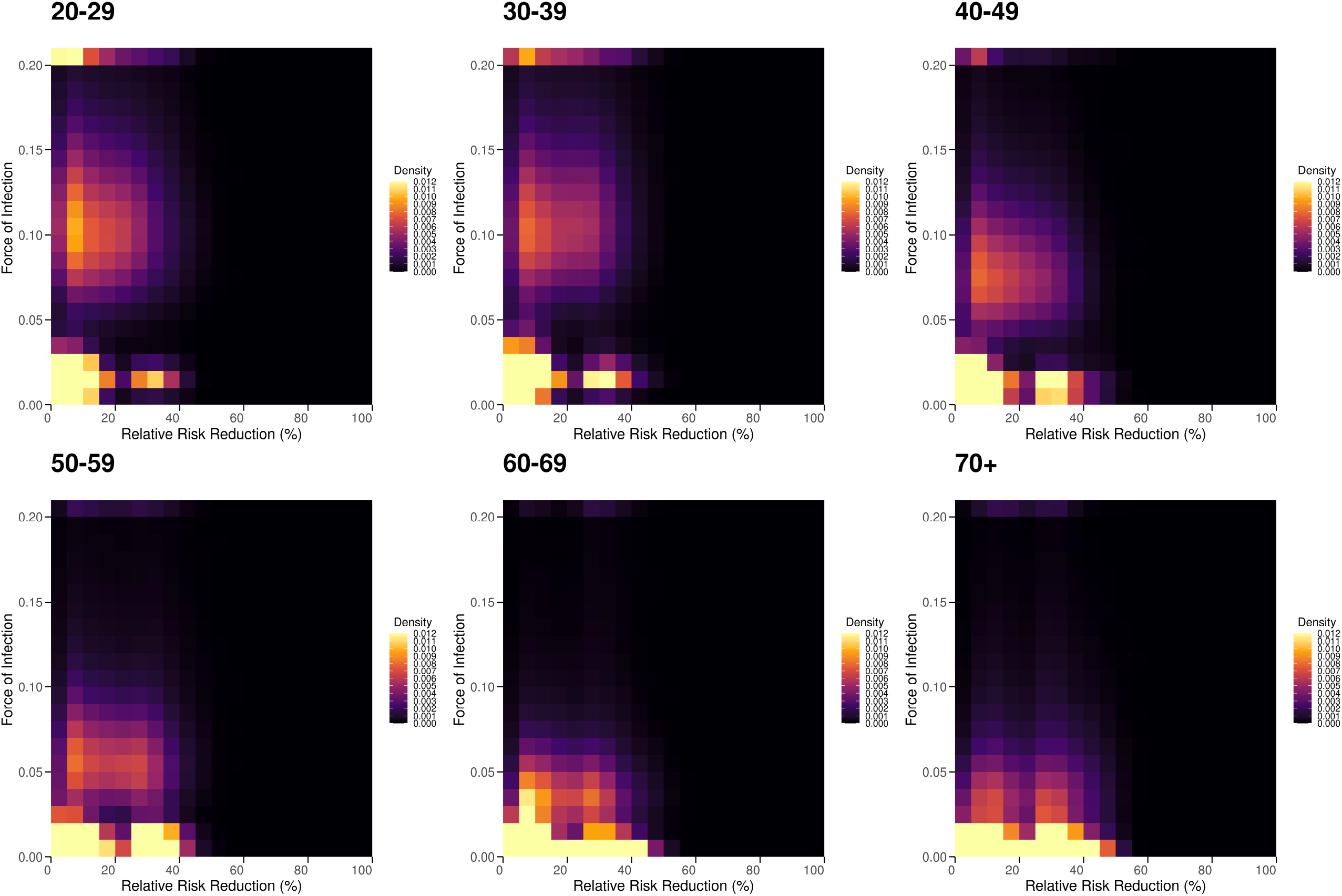
Heatmaps of the distribution of the force of infection and relative risk reduction of infection by age group. In each panel, the cells represent the population fractions within discretized two-dimensional intervals of the force of infection protection level (expressed as the relative risk reduction). The cells with colors closer to yellow represent high density, whereas those closer to black indicate low density.

## 3. DISCUSSION

We proposed a statistical framework based on a questionnaire survey via the Internet to estimate the monthly population-wide incidence of COVID-19, the effect of biological and social immune correlates, and the vulnerability of individuals to COVID-19 based on personal attributes. Importantly, we revealed the inverse relationship between age and FoI after adjustment for the protection offered by acquired immunity and substantial heterogeneity within each age group, as highlighted in the heatmaps in Fig 5. Furthermore, we revealed that personal attributes, including health conditions, are important modulators of the FoI despite most individuals having received vaccination or experienced natural infection at least once.

Several key take-home messages emerged from this study. First, the relatively low protection against JN.1 variants appeared to be induced by vaccination and natural infection. This effect was especially prominent following vaccination with the bivalent Wuhan + Omicron BA. 1/5 vaccine. This result confirms the validity of published studies on JN.1 variants in Japan and aligns with the findings of neutralization studies that suggest substantial immune evasion of JN.1 from immunity provided by exposure to pre-JN.1 infection or vaccines [35–40]. Revealing the mechanism underlying the especially low protection conferred by the bivalent Wuhan + Omicron BA. 1/5 vaccine is beyond the scope of our study; however, our result may be an epidemiological presentation of “immune imprinting” [41–43].

Second, the effects of personal attributes on the risk of infection were also notable and revealed that various underlying health conditions linked with impaired immunity or respiratory or cardiovascular dysfunctions were risk factors for contracting COVID-19. These factors have already been identified as determinants of severe manifestations or death associated with COVID-19 [44–48]. Our finding suggests that, in addition to the importance of vaccinations to curb the risk of developing severe illness by COVID-19, the significance of preventive measures such as wearing masks or avoiding crowded spaces cannot be ignored in individuals with underlying health conditions. In addition to health-related risk, the substantially elevated risk of infection in those with a history of previous infection may reflect unidentified risk factors, including behavioral aspects (e.g., those at risk of death may seek medical attention more often than others) or socioeconomic status, which were not explicitly captured in this study.

Third, another important finding from a population-wide viewpoint is the heterogenous distribution of FoI and protection levels, as highlighted in Figs 3–5. A larger proportion of the younger population was observed in high FoI–low protection regions. By contrast, an opposite pattern was observed in the older population, which was mostly concentrated in low FoI regions with mild heterogeneity, potentially reflecting the recency of infection or vaccination. Because of the difficulty capturing this type of heterogeneity from routine epidemiological surveillance regardless of the disease of interest, our finding is especially important and may guide the effective and efficient implementation of epidemic countermeasures [23]. Furthermore, as post-acute sequelae of COVID-19 gain increasingly more attention, our finding highlights the need to promote vaccination, especially in the working-age population [8,9,49].

Two technical advantages of our framework were identified. First, our findings are based exclusively on a rapidly implementable and simple Internet-based approach that reveals the characteristics of personal immune protection. This point is critical because individual-level profiles regarding immunity against COVID-19 are growing increasingly complex given repeated vaccination and infections and newly emerging SARS-CoV-2 variants. In this context, our framework is expected to be an important addition to an array of conventional approaches that evaluate the effectiveness of vaccine- or infection-induced immunity based on prospective recruitment of study participants over several months or retrospective analysis [21,22,50–52].

Second, our proposed approach of linking the questionnaire respondents to several categories of the census enabled the estimation of both incidence rates and immune protection at the population level. Using our framework to estimate incidence rates may be a complementary approach to epidemiological surveillance, which is essential for tracking key epidemiological indicators, including case-fatality and case-hospitalization ratios. Such an approach is especially useful given that conventional epidemiological surveillance is increasingly limited. Population-level immune protection cannot be estimated with conventional approaches given current data limitations [24,25]. Thus, our framework may be a useful and cost-effective alternative to achieve this aim. Moreover, by widening its scope to infectious diseases other than COVID-19, our framework may guide the implementation of more effective and efficient public health interventions based on risk stratification by age group [23].

Several key limitations of our study should be discussed. First, because the questionnaire data were based on self-reporting by respondents and were not objective data (i.e., as in medical records or serological tests) that are based on medical or technical judgment, the precision of the results may have been limited. As shown in Figs S2 and S3, comparisons between our estimates and limited data from seroepidemiological surveys conducted by Japanese public health agencies suggested some discrepancies, especially regarding the personal history of infection (corresponding to anti-nucleocapsid antibody positivity in seroepidemiological surveys) [53–56]. This gap may have arisen for various reasons, including incorrect answers from the respondents and ascertainment bias. Developing an approach to fill this gap is a future scope of study. Second, small discrepancies between the 2020 census and our estimates of population distribution by sex, age, prefecture, and job category may be attributable to the small sample size or respondent bias in our questionnaire and may have slightly distorted our population-level estimates of incidence rates or protection levels. Our future research scope includes enhancing the robustness of population-level estimates by conducting larger-scale surveys, potentially with the engagement of several survey companies. Finally, our usage of a discrete monthly time scale in the questionnaire survey and the categorization of vaccines and SARS-CoV-2 variants may have been an oversimplification.

In conclusion, we estimated the incidence rate, immune protection of the population, and relative risk of infection attributable to individual factors and converted these estimates to population-level results using our statistical weighting method. Our framework is an important addition to existing epidemiological surveillance programs and studies that focus on vaccine- or infection-induced immunity. Implementing this framework not only would provide a better understanding of epidemiological dynamics but also may enhance our knowledge of how immunological and non-immunological individual-level attributes affect the risk of infection across many infectious diseases and not only COVID-19.

## 4. MATERIALS AND METHODS

### 4.1. Data used in the study

#### 4.1.1. Questionnaire survey

The three key objectives of this study were to 1) estimate the incidence rate of COVID-19 in March 2024 in Japan, 2) characterize the effectiveness of vaccine or infection-induced immunity (peak immunity, decay over time, difference by history of vaccination or infection), and 3) estimate the impact of individual-level attributes including health status on the risk of contracting COVID-19.

Given these objectives and by referring to previously reported information on risk factors for infection and severe outcomes of COVID-19 [44–48], we designed the questionnaire to collect the following information from all respondents:

1. Diagnosis of COVID-19 between 1 and 29 February 2024
2. Biological and health-related background: age, sex, and underlying chronic conditions
3. Social background: profession (occupation), educational status, and household size
4. History of COVID-19 vaccination and natural infection. The details of each questionnaire item can be found in the S1 Text.

An Internet-based questionnaire survey was conducted by a private company (MelLinks Co. Ltd, Tokyo, Japan) in Japan that specializes in Internet-based social surveys using a method that was used in a published study [57]. Among the group of pre-registered monitor respondents that are retained by MelLinks, the recruitment of respondents started on 7 March 2024 and ended on 13 March 2024. The invitation to participate remained open until the distributions of age and prefecture among respondents were proportional to the census data.

The questionnaire was designed to collect information monthly, and therefore we excluded 1) those who had a history of both infection and vaccination in February 2024 and 2) those who reported infection in January 2024, to exclude the potential overlap of the illness period between January and February 2024.

#### 4.1.2. Census data of Japan

To perform the statistical weighting of survey responses, we retrieved the demographic data from the 2020 Population Census of Japan conducted by the Statistics Bureau of the Ministry of Internal Affairs and Communications [58]. We used the following data categories that are stratified by sex, age group, and prefecture:

- Male and female population aged 20 years or older at the national level
- Age distribution by prefecture and sex
- Job category distribution by age and sex in each prefecture

#### 4.1.3. Data on SARS-CoV-2 variants and vaccine types in Japan

We retrieved the historical data on the proportion of SARS-CoV-2 variants in Japan from the National Institute of Infectious Diseases website [59]. We classified the period between January 2020 and January 2024 into the following three antigenically distinct periods for subsequent analyses:

- Pre-Omicron period: January 2020–December 2021
- Pre-XBB Omicron period: January 2022–April 2023
- XBB period: May 2023–December 2023

Notably, SARS-CoV-2 JN.1 variants, the descendants of BA.2.86, were the dominant strains in February 2024.

The types of mRNA vaccines in Japan were classified by period as follows [60–62]:

- Monovalent vaccine adapted to ancestral (Wuhan) strain: February 2021–September 2022
- Bivalent vaccine adapted to ancestral (Wuhan) & Omicron BA. 1/5: October 2022–September 2023
- Monovalent vaccine adapted to XBB.1.5: September 2023–January 2024

For most periods, vaccines manufactured by Pfizer Inc. and Moderna Inc. were available and publicly administered free of charge in Japan for those aged 12 years or older. However, we did not discriminate between these vaccine types to ensure simplicity and because of the practical difficulty in collecting details on vaccine type from all of the respondents.

### 4.2. Statistical Model and Inference in the study

#### 4.2.1. Modeling immune decay after vaccination or infection before January 2024

In analogy to how vaccine effectiveness in an epidemiological sense is generally expressed as 1 − *RR* (*RR*: relative risk), we hereafter use the letter *V* to represent the effectiveness of protection conferred by vaccination or infection. We modeled *V*_*i*_, the protection level against symptomatic infection of an individual *i*, as a function of time since the last immunizing event, which may occur either after vaccination or infection. For those without a history of infection or vaccination, we assumed *V*_*i*_ = 0. Otherwise, we assumed a biexponential waning of protection analogous to the antibody-waning model of Hogan et al.[63] In contrast to Hogan et al., we directly applied the biexponential model to the protection to keep the model simple yet continue to reflect the biological dynamics of antibodies that offer protection. The model for relative risk reduction at calendar time *t* was as follows:

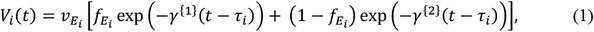

where *E*_*i*_ stands for the last immunizing event of an individual 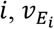 is the peak protection level for *E*_*i*_, and *τ*_*i*_ is the calendar time of the last immunizing event for individual *i*. Both *γ*^{1}^ and *γ*^{2}^ are exponential decay rates that characterize short- and long-lasting protections, and 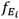 is the fraction of the contribution of short- and long-lasting protections at the peak just after immunization. We assumed that 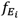 only differs between infection and vaccination, and *γ*^{1}^ and *γ*^{2}^ are fixed parameters regardless of *E*_*i*_.

#### 4.2.1. Modeling of the risk of infection by immunization status and other individual-level factors

First, we assumed that the susceptibility of an individual *i* is purely attributable to a specific (acquired) immunity that can be written as

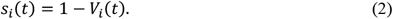

Then, we modeled the FoI for an individual *i* as follows:

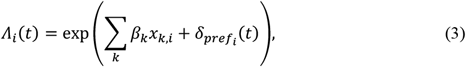

where a linear combination of covariates *x*_*k*,*i*_ is adjusted by prefectural random effects. The first six terms of ∑_*k*_ *β*_*k*_*x*_*k*,*i*_ including the intercept represent the baseline hazard for each age group as

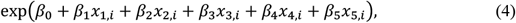

where exp(*β*_0_) is modeled as the baseline hazard of 20–29 years, and *x*_1_~*x*_5_ are dummy variables that represent the age group to which individual *i* belong. The full list of covariates is provided in Table 1.

Upon exposure to Λ_*i*_(*t*), individual *i* experiences the effective hazard *s*_*i*_(*t*)Λ_*i*_(*t*), considering the effect of protective immunity. Thus, the probability of individual *i* being infected from time *t*_1_ to *t*_2_ can be expressed as

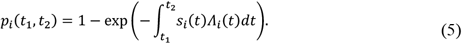

In this study, all of the information from respondents in our dataset was chronologically discretized by month. Moreover, our focus was the occurrence of infection in February 2024. Therefore, assuming that *s*_*i*_ and Λ_*i*_(*t*) were constant throughout February 2024 (i.e., 1 month = *t*_2_-*t*_1_), we simplified the notation of *p*_*i*_(*t*_1_, *t*_2_) as

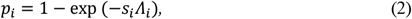

where *s*_*i*_ and Λ_*i*_ represent constant susceptibility and FoI, respectively, for individual *i* throughout February 2024. Using *p*_*i*_, the binomial likelihood for observing the questionnaire-based incidence data *D*^*Q*^ is expressed as follows:

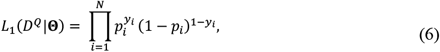

where *y*_*i*_ = 1 (or 0) indicates that individual *i* was infected (or not infected) in February 2024 and Θ = {*v, γ*, *β*, δ}.

#### 4.2.2. Estimation of population-level incidence and immunity

To estimate the population-wide incidence rate and immune profile in the entire Japanese population, the statistical weighting of the results from individuals who responded to our survey was needed. The Japanese 2020 census was used as the reference data for this weighting. The following describes the statistical weighting framework, a conceptual variant of the Raking method, which is used to match survey data from a sampled group to a reference population [64]:

Let 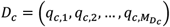 be arbitrary population-wide data, where *c* represents the data category of 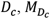 is the number of subcategories in *D*_*c*_, and *q*_*l*_ represents the number of people belonging to subcategory 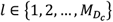. Given that the subset of questionnaire-based data 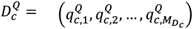 that also represents data category *c*, consider some adjustment or weighting to 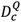 to obtain a dataset comparable to *D*_*c*_. For this aim, we introduced a weighting α = [α_1_, α_2_, …, α_*N*_], ∑ α_*j*_ = 1, where each α_*j*_ is a weight for each individual *j* who responded to the questionnaire. Then, finding the optimal weight α is essentially inferring α based on the likelihood 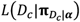 defined by the model

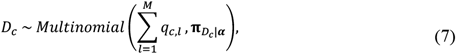

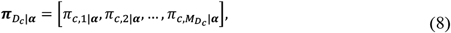

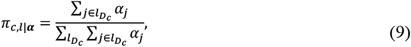

where 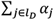 represents the sum of α_*j*_ over all *j* that belong to category *l*_*D*_.

In this study, the three data categories from the 2020 census described above were used for the above-mentioned fitting protocol. By naming the set of these three data categories as *C*_*census*_, the likelihood of observing *D* given α can be written as a combination of multinomial likelihoods:

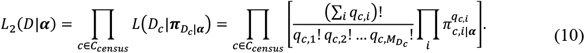

The estimated values of α were used to calculate census-equivalent population-level estimates including incidence rates, FoI, and susceptibility. Briefly, to obtain such an estimate on an arbitrary category *A* that may stand for an estimated value or a binary variable that represents responses to a specific survey question, the group-level estimate of *A* regarding the group of interest *G* can be calculated as a weighted average:

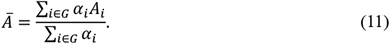

The estimated values of α were also used to obtain the census-equivalent population-level density by FoI and susceptibility level. When considering intervals ***U*** = {*U*_1_, *U*_2_, …} for *A*, the density in an interval *I*_*j*_ can be calculated as

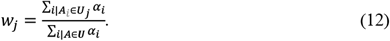

#### 4.2.4. Statistical inference of parameters

We estimated Θ and α using the total likelihood *L* = *L*_1_(*D*^*Q*^|Θ)*L*_2_(*D*|α) of the Markov Chain Monte Carlo method together with the prior distributions and information as follows (see S2 Text for details):

- prior distributions for *β*_*k*≥2_ were designed for the implementation of a Bayesian lasso framework [65]
- weak to non-informative priors were used for other parameters
- findings from published studies on vaccine effectiveness against JN.1 variants were also reflected [35,36,66]

For each of the four Markov Chain Monte Carlo chains, we generated 1,500 samples and discarded the first 500 warmup iterations to generate a total of 1,000 posterior samples. A total of 4,000 posterior samples were collected. We confirmed that R-hat statistics for all parameters were below 1.01 and that all default diagnostics of CmdStan returned no issues regarding convergence. All of the analyses were conducted in R (version 4.2.2) and Stan via CmdStan (version 2.34.0) [67–69].

## Acknowledgments

We thank Anahid Pinchis, MBA, BSc from Edanz (https://jp.edanz.com/ac) for editing a draft of this manuscript.

## Data Availability

The anonymized data from the questionnaire survey in this study cannot be disclosed because of ethical and legal restrictions agreed upon during the data collection process. In lieu of the data from the original questionnaire, two sample data for demonstrative purposes are provided as supplementary data that were synthesized on the basis of Bayesian networks that are estimated from the original data using pgmpy [70]. These sample data are available as S10 Data and S11 Data. the sample codes that can be applied to these sample data can be accessed from https://github.com/pk2393/covid_qs_feb2024. Data from the 2020 census that are used in this study are also provided as supplementary data. The codes used for the analyses in this study are also accessible at https://github.com/pk2393/covid_qs_feb2024.

## Ethical approval statement

The questionnaire survey conducted in this study was approved by the institutional ethics committee of Kyoto University (number R4232). No ethical approval was required for the census data given its openly accessible nature.

## Supporting Information

**S1 Text: Information collected with the questionnaire survey from each respondent in the study**

**S2 Text: Prior distributions and information used for Bayesian inference by Markov Chain Monte Carlo method**

**S1 Table: Descriptive analysis of survey respondents**

**S2 Table: Estimated values of parameters characterizing immune protection dynamics**

**S3 Table. Number of respondents by job category linked to the Census**

**S1 Fig. Posterior estimates of the prefectural effects that modify personal force of infection**.

**S2 Fig. Comparison of weighted estimates of past exposure to COVID-19 infection and vaccination with serological surveys (donated blood** [55,56] **and the residual of clinical serum samples at commercial clinical testing laboratories** [53,54]**) in the Japanese male population in January and March 2024**. For serological surveys, results of anti-nucleocapsid antibody positivity are shown in the “Past Infection” panels, whereas results of anti-spike antibody positivity are shown in the “Any Exposure” panels. *results shown for “70+-year” age group for the residual serum sample survey are actually for the “70-79” age group in the survey because of the difference in age group stratification. [53,54] **no results for the “70+-year” age group and “anti-spike antibody” in the donated blood survey. [55,56]

**S3 Fig. Comparison of weighted estimates on the past exposure to COVID-19 infection and vaccination with serological surveys (donated blood** [55,56] **and the residual of clinical serum samples at commercial clinical testing laboratories** [53,54]**) in the Japanese female population in January and March 2024. For serological surveys, results of anti-nucleocapsid antibody positivity are shown in the “Past Infection” panels, whereas results of anti-spike antibody positivity are shown in the “Any Exposure” panels**.

*results shown for “70+-year” age group for the residual serum sample survey are actually from the “70-79-year” age group in the survey because of the difference in age group stratification. [53,54] **no results for the “70+-year” age group and “anti-spike antibody” in the donated blood survey. [55,56]

S1 Data. Estimated prefecture-wise population distribution by age group, male population

S2 Data. Estimated prefecture-wise population distribution by age group, female population

S3 Data. Estimated job type distribution by age group, male population

S4 Data. Estimated job type distribution by age group, female population S5 Data. Census data of the male and female population by age group

S6 Data. Census data of the male population by age group and prefecture

S7 Data. Census data of the female population by age group and prefecture

S8 Data. Census data of the male population by job category and by age group

S9 Data. Census data of the female population by job category and by age group

S10 Data. Synthetic data for estimation of the effects of personal attributes on the force of infection

S11 Data. Synthetic data for estimation of statistical weighting to fit the respondent cohort to the census data

