## supplementary material for "Reconstructing the force of infection and immune fraction of the population via a single snapshot survey: a case study of COVID-19 in Japan"

**Supplementary Materials**

**Supplementary Methods 1: Information collected with the questionnaire survey from each respondent in the study**

1. Biological sex
2. Age
3. Diagnosis of COVID-19 in Feb 2024
4. Last month of infection (if any)
5. Last month of vaccination (if any)
6. Underlying health conditions
   -Diagnosed with diabetes (regardless of type 1 or type 2).
   -Currently untreated or under treatment for a malignant neoplasm.
   -Under treatment with immunosuppressive drugs, including adrenal corticosteroids, for conditions other than the above.
   -Currently or previously diagnosed with heart disease, such as myocardial infarction, angina, arrhythmia, or heart failure.
   -Currently diagnosed with one of the following: bronchial asthma, chronic obstructive pulmonary disease (COPD), interstitial pneumonia, pulmonary embolism, pulmonary hypertension, or bronchiectasis (excluding asthma limited to childhood).
   -Currently under treatment for or diagnosed with cerebrovascular diseases, such as cerebral infarction or cerebral hemorrhage.
   -Diagnosed with chronic liver diseases, such as liver cirrhosis, fatty liver, alcoholic liver disease, or autoimmune hepatitis.
   -Body Mass Index (BMI) of 30 $kg/m^2$ or higher: BMI = weight (kg) / [height (m) × height (m)].
7. Drinks alcohol at least once a week on average (regardless of the amount).
8. Smoking habit (including occasional smoking, not necessarily daily).
9. Household size (1 or >1)
10. Prefecture of residence
11. Job categories: based on the 2020 Census [1,2]
   -Agriculture, Forestry
   -Fisheries
   -Mining, Quarrying, and Gravel Extraction
   -Construction
   -Manufacturing
   -Electricity, Gas, Heat Supply, and Water Utilities

-Information and Communications
-Transportation and Postal Services
-Wholesale and Retail Trade
-Finance and Insurance
-Real Estate and Goods Rental and Leasing
-Academic Research, Professional, and Technical Services
-Accommodation and Food Services
-Living-Related and Amusement Services
-Education and Learning Support
-Medical and Welfare Services
-Combined Services
-Services (not elsewhere classified)
-Public Administration (not elsewhere classified)
-Industry Not Classifiable
-Unemployed

**Supplementary Methods 2: Prior distributions and information used for Bayesian inference by Markov Chain Monte Carlo method**

1: Immune protection and its decay

$$v_E \sim Beta(10, 10),$$
$$E: all\ types\ of\ exposure. \quad \text{(S1)}$$

$$h^{\{1\}} \left(= \frac{\log(2)}{\gamma^{\{1\}}}\right) \sim Normal(60, 10), \quad \text{(S2)}$$

$$h^{\{2\}} \left(= \frac{\log(2)}{\gamma^{\{2\}}}\right) \sim Normal(360, 120), \quad \text{(S3)}$$

$$f_{infect}, f_{vaccine} \sim Beta(20, 20), \quad \text{(S4)}$$

For $v_E$ and $f$, priors were arbitrarily provided by assuming mild concentrations around 50%. Priors for $h^{\{1\}}$ and $h^{\{2\}}$ were based on those used in Hogan et al [3] with minor modification.

2: Force of Infection

$$\beta_0 \sim Norma\ l(0, 10), \quad \text{(S5)}$$

$$\beta_{i \geq 1} \sim Laplace(0, \tau), \quad \text{(S6)}$$

$$\tau \sim Half\ Cauchy(0, 1), \quad \text{(S7)}$$

$$\delta_{pref_i} \sim Normal(0, \sigma_\delta), \quad \text{(S8)}$$

$$\sigma_\delta \sim Inverse\ Gamma(2, 0.5), \quad \text{(S9)}$$

The prior distribution for $\beta_0$ was arbitrarily set, whereas Laplace distributions as priors for $\beta_{i \geq 1}$ were designed to be defined by hyperparameter $\tau$. Prefectural effects $\delta_{pref_i}$ were assumed to follow normal distributions with standard deviation $\sigma_\delta$, that is also a hyperparameter. Note that a constraint $\sum_i \delta_{pref_i} = 1$ was posed to ensure identifiability of $\delta_{pref_i}$.

3: Weighting

$$\boldsymbol{\alpha} \sim Dirichlet(\boldsymbol{u}), \quad \text{(S10)}$$

$$\boldsymbol{u} = (5, 5, \dots, 5). \quad \text{(S11)}$$

All elements of $\boldsymbol{u}$ (a vector with 7166 elements) were arbitrarily set to 5, leading to the assumption that each value in $\boldsymbol{\alpha}$ has approximately $1\%$ probability of taking values less than $\frac{1}{4\times7166}$, or one-fourth of the expected probability without additional information.

4. Published studies on the protection against JN.1 subvariant infection by vaccine or infection

Using the protection function $R(\Delta\tau)$ in analogy to $V_i(t)$ in equation (1) in the main text:

$$V(\Delta\tau|E) = v_E\left[f_E * \exp\left(-\gamma^{\{1\}}\Delta\tau\right) + (1 - f_E) * \exp\left(-\gamma^{\{2\}}\Delta\tau\right)\right], \quad \text{(S12)}$$

we defined the following likelihoods based on published studies:

a) Kirwan et al. [4]

$$\log\left(1 - V(30|vaccine, XBB)\right) \sim Normal\left(\log(1 - 0.422), \frac{log(1 - 0.217) - log(1 - 0.603)}{3.92}\right), \quad \text{(S13)}$$

$$\log\left(1 - V(90|vaccine, XBB)\right) \sim Normal\left(log(1 - 0.241), \frac{log(1 - 0.007) - log(1 - 0.429)}{3.92}\right), \quad \text{(S14)}$$

$$\log\left(1 - V(150|vaccine, XBB)\right) \sim Normal\left(log(1 - 0.267), \frac{log(1 + 0.275) - log(1 - 0.579)}{3.92}\right), \quad \text{(S15)}$$

$$\log\left(1 - V(30|vaccine, Wuhan + Omicron\ BA.1/5)\right) \sim Normal\left(log(1 - 0.022), \frac{log(1 + 0.357) - log(1 - 0.295)}{3.92}\right), \quad \text{(S16)}$$

$$\log\left(1 - V(90|vaccine, Wuhan + Omicron\ BA.1/5)\right) \sim Normal\left(log(1 - 0.151), \frac{log(1 + 0.554) - log(1 - 0.536)}{3.92}\right), \quad \text{(S17)}$$

$$\log\left(1 - V(90|infection, \quad XBB)\right) \sim Normal\left(log(1 - 0.493), \frac{log(1 - 0.292) - log(1 - 0.636)}{3.92}\right). \quad \text{(S18)}$$

b) Huiberts et al.[5]

$$\log\left(1 - V(30|vaccine, XBB)\right) \sim Normallog(1 - 0.45), \frac{\left(log(1 - 0.3) - log(1 - 0.6)\right)}{3.92}. \quad \text{(S19)}$$

c) Link-Gelles et al.[6]

$$\log\left(1 - \frac{1 - V(80|vaccine, XBB)}{1 - V(674|vaccine, Wuhan + Omicron\ BA.1/5)}\right) \sim Normal\left(log(1 - 0.49), \frac{log(1 - 0.19) - log(1 - 0.68)}{3.92}\right). \tag{S20}$$

**Supplementary Fig 1. Posterior estimates of the prefectural effects that modify personal force of infection**

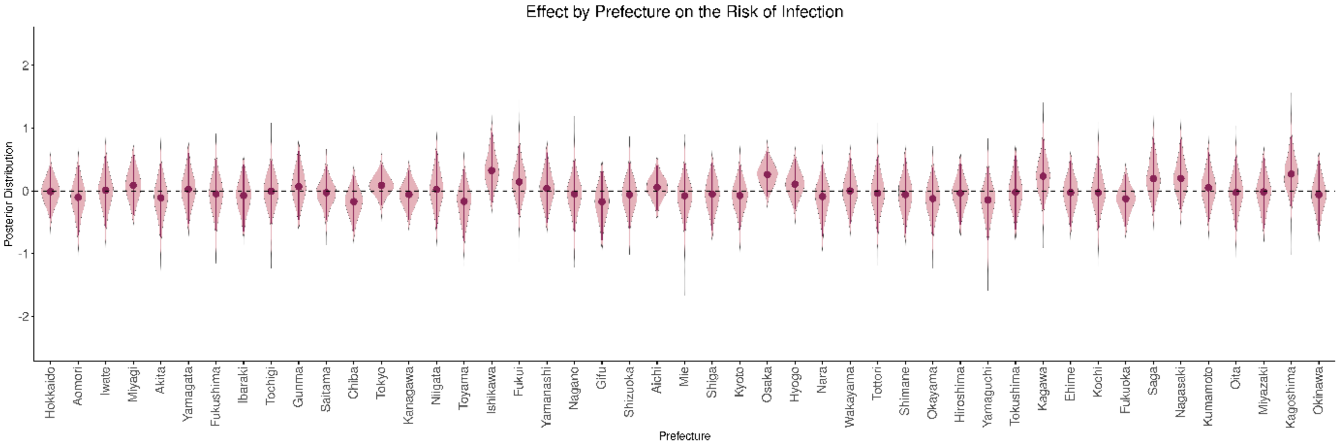

**Supplementary Fig 2. Comparison of weighted estimates of past exposure to COVID-19 infection and vaccination with serological surveys (donated blood** [7,8] **and the residual of clinical serum samples at commercial clinical testing laboratories** [9,10]**) in the Japanese male population in January and March 2024.** For serological surveys, results of anti-nucleocapsid antibody positivity are shown in the "Past Infection" panels, whereas results of anti-spike antibody positivity are shown in the "Any Exposure" panels.

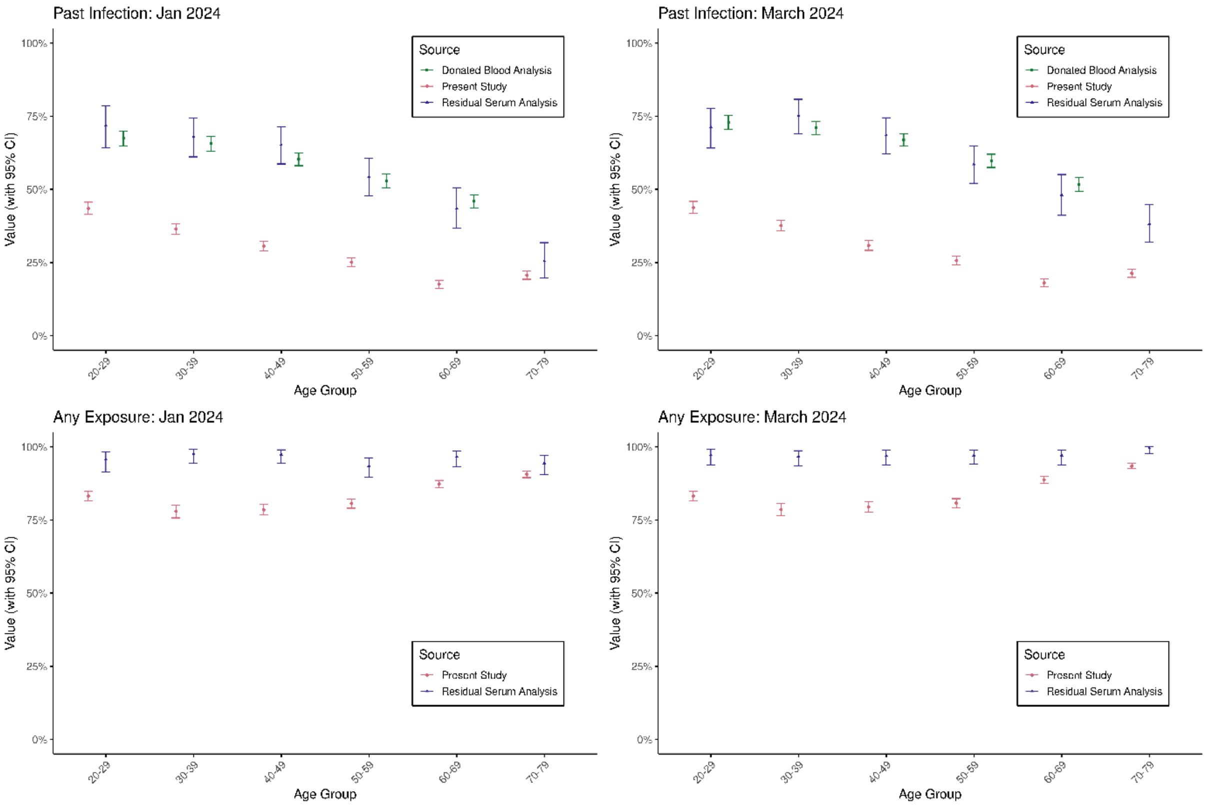


*results shown for "70+-year" age group for the residual serum sample survey are actually for the "70-79" age group in the survey because of the difference in age group stratification. [9,10]

**no results for the "70+-year" age group and "anti-spike antibody" in the donated blood survey. [7,8]

**Supplementary Fig 3. Comparison of weighted estimates on the past exposure to COVID-19 infection and vaccination with serological surveys (donated blood** [7,8] **and the residual of clinical serum samples at commercial clinical testing laboratories** [9,10]**) in the Japanese female population in January and March 2024. For serological surveys, results of anti-nucleocapsid antibody positivity are shown in the "Past Infection" panels, whereas results of anti-spike antibody positivity are shown in the "Any Exposure" panels.**

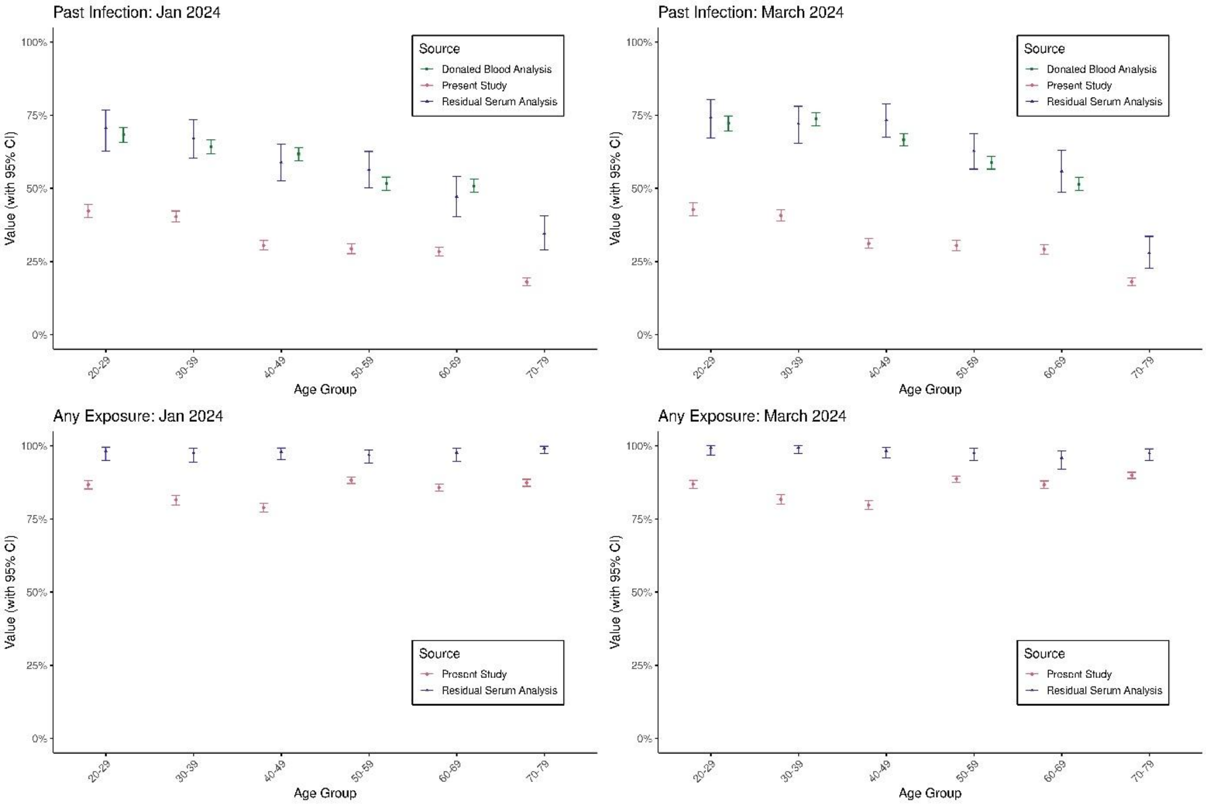


*results shown for "70+-year" age group for the residual serum sample survey are actually from the "70-79-year" age group in the survey because of the difference in age group stratification. [9,10]

**no results for the "70+-year" age group and "anti-spike antibody" in the donated blood survey. [7,8]

**Supplementary Table 1. Descriptive analysis of survey respondents**

| Category | Total | COVID-19 in Feb 2024 | No COVID-19 In Feb 2024 | % diagnosed In Feb 2024 |
|---|---|---|---|---|
| Total | 7116 | 251 | 6915 | 3.53 |
| Age Group (years) | | | | |
| 20-29 | 1196 | 79 | 1117 | 6.61 |
| 30-39 | 1187 | 66 | 1121 | 5.56 |
| 40-49 | 1197 | 41 | 1156 | 3.43 |
| 50-59 | 1190 | 28 | 1162 | 2.35 |
| 60-60 | 1194 | 18 | 1176 | 1.51 |
| 70 and over | 1202 | 19 | 1183 | 1.58 |
| Sex | | | | |
| Male | 3592 | 145 | 3447 | 4.04 |
| Female | 3574 | 106 | 3468 | 2.97 |
| History of Vaccination | | | | |
| No | 1257 | 20 | 1237 | 1.59 |
| Yes | 5678 | 231 | 5678 | 4.07 |
| Last Vaccination | | | | |
| XBB.1.5 | 1373 | 38 | 1335 | 2.77 |
| Wuhan + Omicron BA. 1/5 | 2060 | 56 | 2004 | 2.72 |
| Wuhan | 2476 | 137 | 2339 | 5.53 |
| Infection before Dec 2023 | | | | |
| No | 4867 | 49 | 4818 | 1.01 |
| Yes | 2299 | 202 | 2097 | 8.79 |
| Diabetes Mellitus | 343 | 25 | 318 | 7.29 |
| Neoplastic Disorder | 109 | 25 | 84 | 22.94 |
| Immune Suppression | 91 | 16 | 75 | 17.58 |
| Respiratory Disorder | 219 | 27 | 192 | 12.33 |
| Cardiovascular Disorder | 220 | 21 | 199 | 9.55 |
| Cerebrovascular Disorder | 91 | 10 | 81 | 10.99 |
| Liver Disorder | 100 | 12 | 88 | 12 |
| Obesity (BMI >30kg/$m^2$) | 240 | 10 | 230 | 4.17 |
| Smoking | 866 | 26 | 840 | 3 |
| Drinking | 1293 | 50 | 1243 | 3.87 |

| | | | | |
|---|---|---|---|---|
| Household size | | | | |
| 1 | 1529 | 60 | 1469 | 3.92 |
| More than 1 | 5637 | 191 | 5446 | 3.39 |

**Supplementary Table 2. Estimated values of parameters characterizing immune protection dynamics**

| Parameters | Estimated values (95% CrI) |
|---|---|
| $v_{infect,XBB}$ | 0.555 (0.380, 0.712) |
| $v_{infect,pre-XBB\ Omicron}$ | 0.611 (0.392, 0.797) |
| $v_{infect,pre-Omicron}$ | 0.499 (0.290, 0.705) |
| $v_{vaccine,XBB}$ | 0.548 (0.452, 0.650) |
| $v_{vaccine,Wuhan+\ Omicron\ BA.1/5}$ | 0.347 (0.213, 0.495) |
| $v_{vaccine,Wuhan}$ | 0.441 (0.239, 0.661) |
| $h^{\{1\}}\ \left(=\frac{\log(2)}{\gamma^{\{1\}}}\right)$ (days) | 62.0 (41.7, 81.5) |
| $h^{\{2\}}\ \left(=\frac{\log(2)}{\gamma^{\{2\}}}\right)$ (days) | 439.5 (253.1, 644.3) |
| $f_{infect}$ | 0.412 (0.273, 0.570) |
| $f_{vaccine}$ | 0.524 (0.372, 0.669) |

**Supplementary Table 3. Number of respondents by job category linked to the Census**

| Job Category | |
|---|---|
| Total | 7116 |
| Agriculture, Forestry | 72 |
| Fisheries | 6 |
| Mining, Quarrying, and Gravel Extraction | 15 |
| Construction | 231 |
| Manufacturing | 768 |
| Electricity, Gas, Heat Supply, and Water Utilities | 59 |
| Information and Communications | 217 |
| Transportation and Postal Services | 295 |
| Wholesale and Retail Trade | 504 |
| Finance and Insurance | 174 |
| Real Estate and Goods Rental and Leasing | 125 |
| Academic Research, Professional, and Technical Services | 91 |
| Accommodation and Food Services | 183 |
| Living-Related and Amusement Services | 114 |
| Education and Learning Support | 292 |
| Medical and Welfare Services | 605 |
| Combined Services | 55 |
| Services (not elsewhere classified) | 483 |
| Public Administration (not elsewhere classified) | 270 |
| Industry Not Classifiable | 214 |
| Unemployed | 2393 |
